# The Deep Brain Stimulation Response Network in Parkinson’s Disease Operates in the High Beta Band

**DOI:** 10.1101/2025.04.07.25325381

**Authors:** Bahne H. Bahners, Lukas L. Goede, Patricia Zvarova, Garance M. Meyer, Konstantin Butenko, Roxanne Lofredi, Nanditha Rajamani, Frederic L.W.V.J. Schaper, Clemens Neudorfer, Barbara Hollunder, Julianna Pijar, Savir Madan, Lauren A. Hart, Matthias Sure, Alexandra Steina, Fayed Rassoulou, Christian J. Hartmann, Markus Butz, Jan Hirschmann, Jan Vesper, Katharina Faust, Gerd-Helge Schneider, Tilmann Sander, Michael D. Fox, Kai J. Miller, Alfons Schnitzler, Andrea A. Kühn, Esther Florin, Andreas Horn

**Affiliations:** Center for Brain Circuit Therapeutics, Department of Neurology, Brigham & Women’s Hospital, Harvard Medical School, Boston, MA, USA; Institute of Clinical Neuroscience and Medical Psychology, Medical Faculty and University Hospital Düsseldorf, Heinrich Heine University Düsseldorf, Germany; Department of Neurology, Center for Movement Disorders and Neuromodulation, Medical Faculty and University Hospital Düsseldorf, Heinrich Heine University Düsseldorf, Germany; Movement Disorders and Neuromodulation Unit, Department of Neurology, Charité – Universitätsmedizin Berlin, corporate member of Freie Universität Berlin and Humboldt-Universität zu Berlin, Berlin, Germany; Einstein Center for Neurosciences Berlin, Charité – Universitätsmedizin Berlin, corporate member of Freie Universität Berlin and Humboldt-Universität zu Berlin, Berlin, Germany; Berlin Institute of Health (BIH), Berlin, Germany; Department of Neurosurgery, Massachusetts General Hospital, Harvard Medical School, Boston, MA, USA; Berlin School of Mind and Brain, Humboldt-Universität zu Berlin, Berlin, Germany; Department of Functional Neurosurgery and Stereotaxy, Medical Faculty and University Hospital Düsseldorf, Heinrich Heine University Düsseldorf, Germany; Department of Neurosurgery, Medical Faculty and University Hospital Düsseldorf, Heinrich Heine University Düsseldorf, Germany; Department of Neurosurgery, Charité – Universitätsmedizin Berlin, corporate member of Freie Universität Berlin and Humboldt-Universität zu Berlin, Berlin, Germany; Physikalisch-Technische Bundesanstalt, Abbestraße 2-12, 10587 Berlin, Germany; Department of Physiology and Biomedical Engineering, Mayo Clinic, Rochester, MN, USA; Neurosurgery, Mayo Clinic, Rochester, MN, USA; NeuroCure, Charité – Universitätsmedizin Berlin, corporate member of Freie Universität Berlin and Humboldt-Universität zu Berlin, Berlin, Germany; Deutsches Zentrum für Neurodegenerative Erkrankungen (DZNE), Berlin, Germany

**Keywords:** neurophysiology, high beta, magnetoencephalography

## Abstract

Deep brain stimulation (DBS) of the subthalamic nucleus (STN) improves motor symptoms in patients with Parkinson’s disease. Using functional MRI, optimal DBS response networks have been characterized. However, neural activity associated with Parkinsonian symptoms is magnitudes faster than what can be resolved by this method. While both *spatial* and *temporal* domains of these networks appear critical, no single study has yet investigated both domains simultaneously.

Here, we aim to close this gap using subthalamic local field potentials that were concurrently recorded alongside whole-brain magnetoencephalography in a multi-center cohort of patients that underwent STN-DBS for the treatment of Parkinson’s disease (N = 100 hemispheres). In every cortical vertex, cortico-subthalamic coupling was correlated with stimulation outcomes.

This network spatially resembled fMRI-based findings (R = 0.40, P = 0.039) and explained significant amounts of variance in clinical outcomes (β_std_ = 0.30, P = 0.002), while theta-alpha and low beta coupling did not show significant associations with DBS response (theta-alpha: β_std_ = −0.02, P = 0.805; low beta: β_std_ = −0.08, P = 0.426). The ‘optimal’ high beta coupling map was robust when subjected to various cross-validation designs (10-fold cross-validation: R = 0.29, P = 0.009; split-half design: R = 0.31, P = 0.026) and was able to predict outcomes across DBS centers (R = 0.74; P_(1)_ = 8.9e-5).

We identified a DBS response network that i) resembles the previously defined MRI network and ii) operates in the high-beta band. Maximal connectivity to this network was associated with optimal DBS outcomes and was able to cross-predict clinical improvements across DBS surgeons and centers.

## Introduction

Deep brain stimulation (DBS) of the subthalamic nucleus (STN) is an effective surgical treatment for Parkinson’s disease^1^. Stimulation of the STN significantly improves motor symptoms and quality of life, and this effect critically depends on the site of stimulation, i.e. electrode placement and DBS programming^1,2^. This has inspired studies that mapped optimal stimulation sites – known as ‘sweet spots’ – onto specific subregions of the nucleus^3,4^. However, from its inception, it has been widely acknowledged that DBS exerts its effects onto distributed *brain networks*, not just onto localized stimulation targets^5,6^. Hence, an optimal network – above and beyond an optimal target site – may represent the true guidance needed to optimize DBS^7^.

Indeed, multiple studies have described the optimal *spatial* topography of a DBS treatment network in Parkinson’s disease^8–12^. Independently, numerous electrophysiological studies have described *temporal* properties of the Parkinsonian network, predominantly as synchronized activity in the beta band^13–20^, and more recently, as circuit activity in the high beta band^14,21^. However, none of these studies has yet explicitly *linked* the spatial and temporal domains of the DBS response network, let alone analyzed the spatial and temporal properties of the network simultaneously.

In the *spatial* domain, studies within the emerging field of ‘connectomic DBS’ have attempted to identify the optimal DBS response network for Parkinson’s disease – most often using noninvasive MRI-based measures (for a review, see^22^). This framework, termed DBS network mapping, has enabled the identification of optimal connectivity profiles whose modulation is associated with maximal symptom improvements. This strand of research led to the characterization of a functional *DBS response network* in Parkinson’s disease ^8,23^, which was reproducible across centers^8^, across DBS targets, such as the STN and GPi^10^, and even across brain stimulation modalities^24^.

On the *temporal* scale, a large body of literature has associated increased oscillatory activity in the *low* beta band (13-20 Hz) acquired from STN local field potential (LFP) recordings with the severity of akinetic-rigid symptoms in Parkinson’s disease ^25–28^. Conversely, activity in cortico-subthalamic networks has been described to be dominated by synchrony in the *high* beta band (21–35 Hz) and to originate from motor and premotor areas^13–17^. Stimulation of the STN has been shown to both attenuate beta power within the STN as well as to disrupt (high) beta coherence between cortex and STN^13,21,27^.

Pooling the *spatial* and *temporal* characteristics of a Parkinson’s disease *treatment* network together, we would hypothesize to identify a polysynaptic network that i) involves the STN and specific premotor regions and ii) communicates in the high beta band. Despite these independent spatial or temporal characterizations, no prior study has analyzed both aspects simultaneously to characterize the Parkinson’s disease response network in both space and time. This is due to limitations of the applied methods, with resting state fMRI resolving very slow temporal scales (network activity in the 0.01-0.1 Hz range)^29,30^, and local field potential recordings being limited to the recording site(s), i.e. not resolving whole-brain topographies.

One framework capable of analyzing networks in both spatial and temporal domains is Magneto-/Electroencephalography (M/EEG)^31,32^. While MEG is uniquely suited to investigate temporal dynamics alongside precise spatial resolution of the cortical convexity, reconstruction of *subcortical* signals is still problematic even with this method. A very promising, but technically elaborate, extension to this method is to amend subcortical MEG data by simultaneously acquired LFP recordings from implanted DBS electrodes^33^. Simultaneous MEG-LFP recordings typically require externalizing DBS electrode extensions between the first (implantation of electrodes) and second surgery (implantation of pulse generator). Because of these hurdles involved, such data has only been measured by a very select number of centers world-wide^16,18–20,33^.

In the present study, we have obtained and reanalyzed a substantial portion of the globally available data of this type (N = 50 patients; N = 100 hemispheres). Using this data, we map the DBS response network for Parkinson’s disease with high spatial and temporal precision.

## Materials and methods

### Patient recruitment and recording procedure

Combined bilateral STN LFP and MEG resting state recordings were available in 86 patients with Parkinson’s disease at two German DBS centers in Düsseldorf and Berlin. MEG-LFP data from 57 patients had previously been analyzed and published^17–19^. From the whole cohort, we retrospectively selected a total of 50 patients (40 from Düsseldorf and 10 from Berlin) for which UPDRS-III hemiscores were available in the medication off state in both stimulation ON and OFF states one year after DBS surgery (see supplementary table 1 for details). All patients underwent a two-staged approach to bilateral STN-DBS surgery. Following implantation, DBS electrode extension wires were externalized at the scalp level to allow connection to an external recording device. Simultaneous LFP-MEG resting state recordings were acquired on the day after surgery for a duration of at least three minutes (range: three to ten minutes of recording time), after a complete overnight withdrawal of dopaminergic medication. Importantly, no stimulation was delivered in this period of time or during recordings (Figure 1). Patients were instructed to keep their eyes open throughout the recording and hold their heads still in the MEG helmet. MEG recordings were acquired using a 306 channel-MEG system (Neuromag, MEGIN Oy, Espoo, Finland) at the Institute of Clinical Neuroscience and Medical Psychology in Düsseldorf and a 125-channel-MEG system (Yokogawa ET 160, Tokyo, Japan) at the Physikalisch-Technische Bundesanstalt in Berlin. The data were recorded with a sampling rate of 2kHz and LFP recordings were obtained simultaneously with MEG by connecting the externalized extensions from the DBS leads to an EEG amplifier which was synchronized with the MEG system and LFP signals were recorded with a mastoid electrode (Düsseldorf) or the uppermost contact of the right DBS electrode (Berlin) as reference. The DBS electrodes used for implantation are featured in supplementary table 1 and included 4- and 8-contact electrode models. Throughout recordings heart beats and eye blinks were monitored using electrocardiography (ECG) and electrooculography (EOG) using the same EEG amplifier. Following the MEG-LFP recording session, DBS electrodes were connected to a pulse generator, that was implanted in a pectoral or abdominal subcutaneous pouch in a second surgical procedure (see Supplementary Table 1 for electrode models). The experimental protocols were approved by the Ethics Committees of the Medical Faculty of Heinrich Heine University Düsseldorf and Charité – Universitätsmedizin Berlin, Campus Virchow Klinikum. The studies were conducted in accordance with the Declaration of Helsinki^40^.

**Figure 1.**
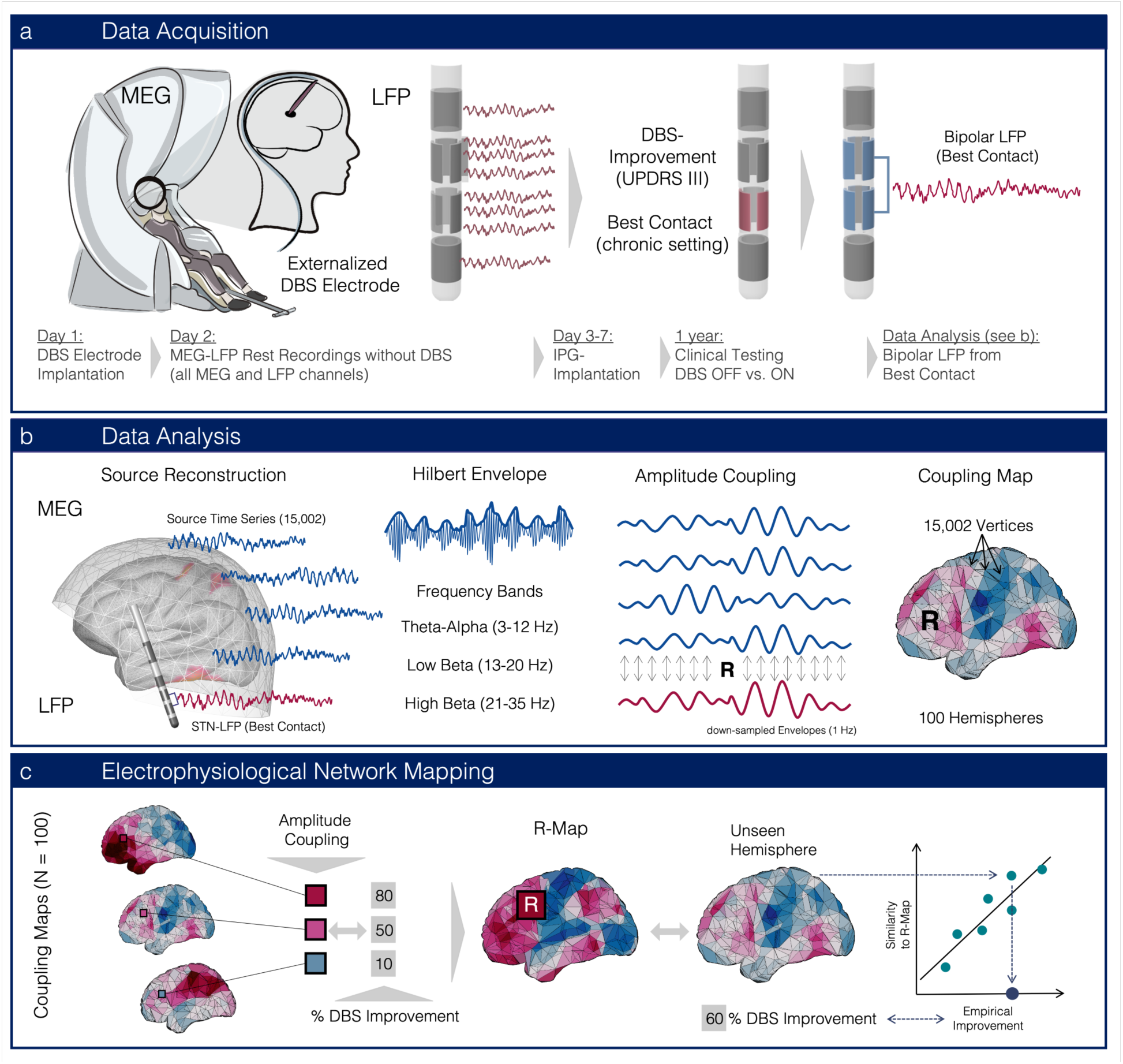
Data analysis and network mapping approach. **a** Magnetoencephalography (MEG) and local field potential (LFP) recordings were simultaneously acquired at rest one day after DBS surgery via externalized DBS lead extensions. All LFP channels were recorded in a monopolar montage (see methods). One year after surgery, the stimulation effect of the chronic DBS setting active at that visit was evaluated using the Unified Parkinson’s Disease Rating Scale part III (UPDRS-III). **b** Cortical source time series were reconstructed from the MEG recordings. LFP were bipolar re-referenced between the clinical DBS contact (used for chronic stimulation at one year) and the adjacent DBS contact. To capture amplitude fluctuations in a particular frequency band over time, Hilbert envelopes were computed in three frequency bands. The resulting amplitude envelopes were down-sampled to 1 Hz^34^ and correlations between the subthalamic (bipolar LFP) and each cortical (source-localized MEG) signal were calculated. This generated an STN coupling map for each of the 100 hemispheres. **c** Coupling maps were correlated vertex-wise with contralateral motor symptom improvements. The resulting correlation map (R-map) represents a connectivity profile associated with optimal symptom improvements. This map may be calculated across all hemispheres, but the process may also be iteratively repeated after leaving out a set of hemispheres (e.g. calculating across 100-x hemispheres). Coupling maps of the x unseen hemispheres were then compared with the ‘optimal’ R-map model by spatial correlations across vertices to predict therapeutic outcomes. According to the model, the more similar the resulting coefficient was, the more ‘optimal’ the electrode in the left-out hemisphere, i.e. the better clinical improvements would be predicted to be. The validity of this model was then tested by correlating similarity coefficients with empirical motor improvements of unseen hemispheres.

### DBS electrode localization

DBS electrodes were localized using Lead-DBS v3.0 for all patients as previously described^35,36^. In brief, postoperative CT and preoperative MRI were co-registered and non-linearly warped to ICBM 2009b Nonlinear Asymmetric (‘MNI’) space using Advanced Normalization Tools (ANTs, https://stnava.github.io/ANTs/). Following brain shift correction due to possible pneumocephalus, the non-linear warp was manually refined using WarpDrive with a focus on the STN region as the target region^37^. Then, electrode trajectories were reconstructed using PaCER^38^ in case of postoperative CT images or TRAC/CORE^39^, if postoperative MRI scans were available.

### LFP & MEG Data analysis

LFP and MEG data were analyzed using Brainstorm^41^. LFP and MEG data were visually screened and epochs containing artifacts as well as bad channels were discarded. The signals were down-sampled to 250 Hz and a notch filter was applied to reduce line noise at 50 Hz and its harmonics up to 250 Hz. The LFP signal was re-referenced to the common average and a high-pass filter at 1 Hz was applied to both, LFP and MEG data. Signal space projection was used to eliminate blink and cardiac artefacts in the MEG data that were detected using the EOG and ECG recordings. The LFP was bipolar re-referenced between the clinically selected DBS electrode one year after surgery and the upper adjacent contact. This selection was adapted to the lower adjacent contact in case it included bad LFP channels, i.e. channels with strong noise or a flat signal or if the uppermost contact of the electrode had been used for chronic stimulation.

Cortical surfaces were reconstructed from preoperative individual T1 MRI using CAT12^42^ and co-registered with the MEG recordings using an iterative closest-point rigid-body registration in Brainstorm. Next, the forward problem was solved with an overlapping spheres head model and 15,002 cortical sources were reconstructed with a linearly constrained minimum variance (LCMV) beamformer^33,43^. For combined MEG-LFP recordings, this method has proven useful to further reduce noise related to the movement of extension cables from DBS electrode to amplifier^33^.

Next, we calculated the Hilbert envelope for the data from all 15,002 reconstructed cortical sources and the bipolar LFP in three frequency bands: theta-alpha (3-12 Hz), low beta (13-20 Hz) and high beta (21-35 Hz)^44,45^. Based on previous work, we applied a wide band infinite impulse response (IIR) filter to obtain the Hilbert envelope^34^. After down-sampling all computed Hilbert envelopes to 1 Hz, individual cortically constrained data were spatially smoothed with a 6 mm Gaussian Kernel^34^. All individual source-level amplitude envelope maps were projected to the Montreal Neurological Institute (MNI) space (ICBM152 2009c Nonlinear Symmetric)^46^ using Shepard’s method as implemented in Brainstorm. Next, the correlation between the STN bipolar LFP amplitude envelope and every cortical amplitude envelope time series was computed (Figure 1). The correlation was computed for both STN-LFP envelope time series separately, resulting in two amplitude coupling maps per patient and frequency band.

We purposefully decided to compute functional connectivity using *amplitude* rather than phase coupling measures (i.e. coherence). When extracting resting state networks from electrophysiological data, phase coupling networks were shown to be less robust in their spatial structure than amplitude coupling networks (i.e. envelope correlation), when spatially compared to resting state fMRI^34,45,48^. Moreover, the role of coherence as a meaningful reflection of functional connectivity (communication-through-coherence hypothesis) has recently been challenged^49,50^.

To enable whole cortex analysis across all 100 hemispheres, we mirrored the vertex values from the left hemisphere to the right hemisphere and vice versa using a symmetric grid derived from the symmetric MNI template (ICBM152 2009c Nonlinear Symmetric). Since no symmetric MNI template was available in Brainstorm yet, we custom-created a novel brainstorm-compatible template with symmetrical cortical vertices which we made publicly available (private link will be updated to DOI for publication: https://figshare.com/s/e4663ae98496ff896fe7). In order to obtain a symmetric grid that exhibited an exact spatial correspondence between left and right vertex coordinates, we reconstructed the cortical surface from the symmetric MNI template using CAT 12 and flipped vertices from the left hemisphere to the right side (x-coordinates from the left were multiplied by −1 in MNI space). Finally, given the differences in recording durations of different experimental protocols (3-10 minutes) and since our analysis focused on the cortical *pattern* of amplitude coupling rather than on absolute coupling values, correlation maps were normalized by dividing all cortical correlation values by their average (mean across vertices) ^15^.

### Electrophysiological network mapping

We gathered the hemisphere-wise percentage improvements in UPDRS-III scores in the medication OFF state one year after surgery (Med OFF / Stim OFF vs. Med OFF / Stim ON) for each of the 100 hemispheres. We also used symptom-specific sub-scores. Importantly, we discarded hemispheres with a baseline score of less than 2 points for the respective symptoms as previously described^11^. This led to smaller group sizes for rigidity (49 hemispheres excluded) and tremor outcomes (58 hemispheres excluded). Next, after mirroring the amplitude coupling maps for 100 hemispheres, in each of the 15,002 cortical vertices we correlated the 100 (hemisphere-wise) amplitude coupling values with the 100 contralateral hemiscore percentage improvement values across the cohort (Figure 1b) to build a mass-univariate model as previously described^8^. This resulted in a correlation map (R-map) which consisted of correlation coefficients that denoted the relationships between amplitude coupling coefficients and DBS outcomes across the cohort for each of the 15,002 vertices.

### Model validation

To validate the intrinsic consistency of the model, we subjected the R-map model to various cross-validation designs. To do so, the R-map was recalculated iteratively, each time leaving out one hemisphere, patient, or fold of hemispheres. Amplitude coupling maps of left out hemispheres were then spatially correlated with the respective R-map that had been derived on the remaining sample (Figure 1). In alignment with previous approaches from the MRI-DBS literature^8^, we hypothesized that the R-map represents an optimal map of amplitude coupling that is associated with maximal DBS improvements. If this hypothesis holds true, spatial similarities between left-out hemispheres and the R-map would be able to estimate variance in DBS improvements. Hence, these spatial similarity coefficients were correlated with the empirical DBS improvements.

As strongly suggested by prior literature findings (see introduction), we hypothesized that high-beta coupling would lead to clinically informative maps. To test this hypothesis, we first fitted a linear model with hemiscore improvement as dependent and spatial similarities of theta-alpha, low beta and high beta coupling maps to the respective R-maps as independent variables. Additionally, we subjected the correlation coefficients between spatial similarities in each frequency band with hemiscore improvements to Fisher-tests with dependent samples to test if the associations between high beta coupling patterns and DBS improvements were significantly stronger than for the other frequency bands. All P-values reported were Bonferroni corrected for multiple comparisons (for the three frequency bands tested).

The R-map was subjected to various cross-validation designs. As a test of superior-most robustness among all of the herein applied approaches, we also included a ‘split-half’ (2-fold) analysis. To achieve this, we created test and training subsets (25 patients each) that balanced the cohorts for baseline symptom severity, improvement and age. We further assessed if specific symptoms would drive the relationship between high beta coupling pattern and DBS outcome and therefore built R-maps using the symptom-specific UPDRS-III sub-scores (see above). These maps were subjected to leave-one-out cross-validation and P-values were again Bonferroni corrected. All analyses were carried out using a two-tailed correlation test (Spearman’s rho).

In a final validation analysis, to rule out center-specific bias, we calculated the total UPDRS-III R-map derived from the Düsseldorf dataset (n = 80 hemispheres) to cross-estimate outcomes in patients operated in Berlin (n = 20 hemispheres). Given that for the cross-prediction between centers only positive correlations are meaningful, a one-sided (right-tailed) significance test was applied for correlation^24^.

Finally, we compared our results with a published fMRI R-map for total UPDRS-III improvement^8^, which is openly available within the Lead-DBS software package. To do so, cortical vertex values were extracted from the whole brain fMRI R-map and spatially rank correlated with the electrophysiological R-map. To test significance of similarities between the two maps against a null-model, we adopted a recently introduced permutation based strategy^47^. Namely DBS outcomes used to calculate the electrophysiological R-map were permuted 10,000 times before iteratively re-calculating the map. Each permuted map was spatially correlated with the fMRI R-map to establish the null model. The spatial correlation coefficient of the unpermuted map was then compared to the distribution of correlation coefficients from permuted maps to assess significance.

### Data availability

The patient imaging and electrophysiology data cannot be publicly shared, since this would compromise patient privacy according to current data protection regulations. They are, however, available from the principal investigators of the collecting sites upon reasonable request within the framework of a data-sharing agreement and the respective local ethical regulations. Inquiries for further information and data-sharing requests should be directed to the corresponding authors of this manuscript (AH, or BHB) who commit to replying to any request within a timeframe of 30 d.

### Code availability

All code used in the analyses presented in this work is openly available within the Lead-DBS environment (https://github.com/leaddbs/leaddbs).

## Results

### Patient demographics and clinical results

50 patients (17 female; mean age: 60.3 ± 9 years) with Parkinson’s disease were included, each having undergone two-stage bilateral STN-DBS implantation at one of two German DBS centers, Düsseldorf (n = 40) or Berlin (n = 10). Motor symptoms, as measured by the motor part of the Unified Parkinson’s Disease Rating Scale (UPDRS-III), significantly responded to stimulation at the follow-up visit one year after surgery in the medication OFF state (stimulation OFF: 41.7 ± 15.3 vs. stimulation ON: 24.4 ± 11 points; t (98) = 6.5, P = 3.4e-9). Importantly, no stimulation or medication was delivered during MEG-LFP recordings and these scores correspond to the one-year follow-up visit (Figure 1). A comprehensive summary of stimulation settings, electrode models and clinical outcomes is provided in supplementary table S1. Demographic data for both cohorts is summarized in supplementary table S2.

### MEG-LFP recordings reveal cortico-STN amplitude coupling dynamics

To measure functional connectivity between the LFP at the chronic DBS contact and all 15,002 cortical sources, cortico-subthalamic amplitude coupling was derived after filtering the signal in three canonical frequency bands that have been shown to be relevant for Parkinsonian pathophysiology: theta-alpha (3-12 Hz), low beta (13-20 Hz) and high beta (21-35 Hz). We hypothesized that – specifically – the high beta coupling signature would show a relevant association with DBS improvements, given that prior literature findings suggested high beta as the dominant frequency band for Parkinsonian *network pathophysiology*^13–17^.

Figure 2 shows the grand average cortico-STN amplitude coupling maps. While theta-alpha coupling consisted in a pattern dominated by frontal sources with a peak in the middle frontal gyrus, low and high beta coupling mapped to Rolandic areas, with low beta showing a more lateral distribution and high beta dominated by mesial cortico-STN coupling in the motor cortex (Figure 2 b-d). Maps were similar across the three frequency bands and confirmed a strong connection between the frontal cortex and STN, with lower frequencies including more associative-limbic domains while faster frequencies more strongly mapping to motor regions^51,52^.

**Figure 2.**
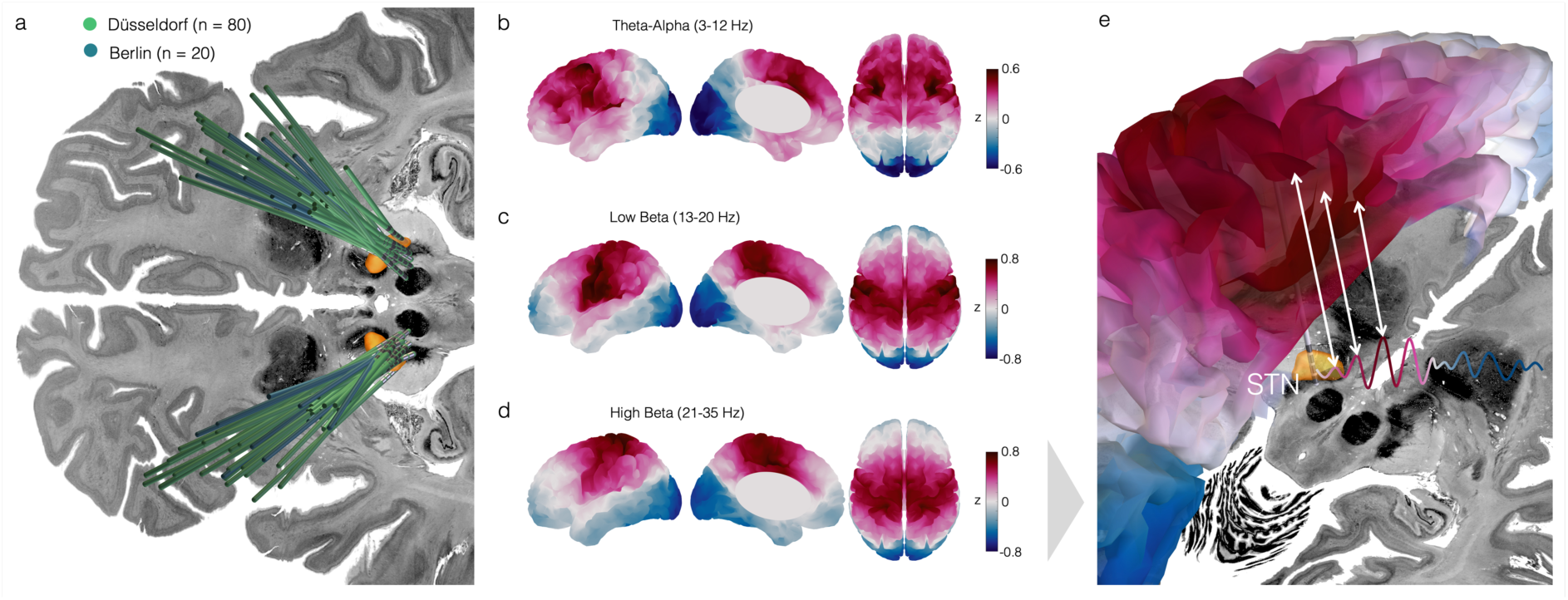
Grand average cortico-subthalamic coupling maps. **a** DBS electrode localizations for 100 STN stimulation sites marked in green (Düsseldorf) and blue (Berlin). The Big Brain and DISTAL atlases were used as a backdrop image and for STN rendering respectively^53,54^. **b-d** Grand average maps (N = 100) of cortico-subthalamic amplitude coupling in three canonical frequency bands between STN time series and the respective ipsilateral hemisphere. Individual STN-cortex amplitude coupling maps in each hemisphere were smoothed, projected to a symmetric MNI template, mirrored for further analysis, then z-scored and averaged across hemispheres for visualization. The theta-alpha coupling map revealed a coupling pattern dominated by frontal regions with a peak in the middle frontal gyrus. Low and high beta coupling were more pronounced in Rolandic areas with a peak in lateral motor areas for low and mesial motor areas for high beta coupling. **e** 3d-rendering of cortical high beta coupling pattern (also shown in panel d) to the subthalamic nucleus (STN, orange) envelope time series (schematic representation).

### High beta coupling signature explains variance in DBS outcomes

Next, we aimed to assess whether the pattern of cortico-STN amplitude coupling was useful in estimating DBS outcomes. Analogously to fMRI-based DBS network mapping^8^, we correlated the amplitude-coupling values in each cortical vertex with the individual DBS motor improvements in the contralateral body (percentage change of contralateral UPDRS-III hemiscores) across the whole cohort (Figure 1 c). This resulted in a correlation map (R-map), which represented the coupling patterns of DBS electrodes associated with maximal symptom improvements. The theta-alpha R-map was dominated by positive correlations in parieto-occipital regions, the low beta R-map revealed a more scattered pattern with positive correlations in frontal and parietal areas (see supplementary Figure S1). The high beta R-map was dominated by positive correlations in frontal regions, while negative correlations mapped to central and occipital areas (Figure 3 a).

**Figure 3.**
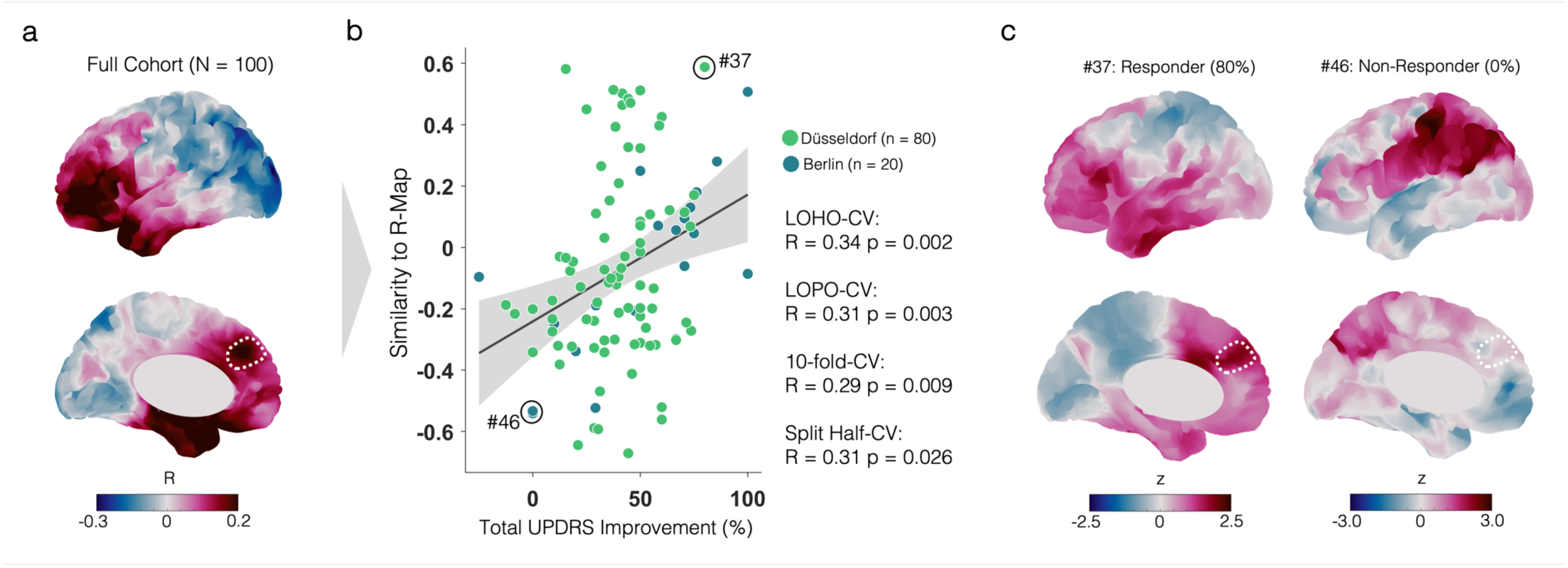
R-map and cross-validations. **a** Correlation map (R-map) generated from high beta coupling maps across the cohort (N = 100 hemispheres). The dashed circle indicates a positive correlation peak observed in the responder map (see C). **b** Correlation plot showing the leave-one-hemisphere-out results. Grey shaded areas represent 95% confidence intervals. Leaving out one hemisphere (leave one hemisphere out cross-validation, LOHO-CV), one patient (leave one patient out, LOPO) or one fold of hemispheres (10-fold cross-validation), the R-map was computed and spatially correlated with the respective unseen map(s) to estimate improvements based on the similarity to the R-map. These estimates were correlated with the actual (empirical) stimulation improvements in total UPDRS scores. In an additional analysis the cohort was split into two subcohorts that were matched by baseline UPDRS, improvement as well as age and a model was built on one half to estimate outcomes in the other half (split half cross-validation). **c** Example coupling maps in the high beta band of two individual patients for one respective hemisphere with a good response (80% for patient #37) or no response (patient #46) are shown.

Next, we built the R-maps for each frequency band again, each time leaving out one hemisphere and spatially correlating this hemisphere’s coupling pattern to the R-map (spatial similarity, see figure 1c). To test our a priori hypothesis that high beta signatures would specifically reflect DBS improvement (more than the lower frequency bands), we fitted a linear model relating hemiscore improvements to theta-alpha, low beta and high beta spatial similarities to the respective R-maps (as independent variables). In this model, the similarities of high beta coupling signatures to the R-map explained significant amounts of variance (β_std_ = 0.30, P = 0.002; here we report standardized beta coefficients, β_std_), while theta-alpha and low beta coupling did not show a significant association (theta-alpha: β_std_ = −0.02, P = 0.805; low beta: β_std_ = −0.08, P = 0.426). Obviously, when comparing the correlation coefficients between similarities to the R-map and hemiscore improvement across the three frequency bands using a Fisher-Test for dependent samples, the association with high beta signatures was significantly stronger than for theta-alpha or low beta (theta-alpha vs. high beta: z = −3.63, P = 2.9e-4; low beta vs. high beta: z = −4.47, P = 8.0e-6). Since the high beta R-map appeared to be most relevant for DBS improvement, we subsequently focused on the validity of this particular map (maps and cross-validations for low beta and theta-alpha bands shown in supplementary figure S1). Despite this, we still applied Bonferroni correction to correct P-values for 3 frequency bands tested.

The high beta R-map was robust when subjected to various cross-validation designs (leave-one-hemisphere out: R = 0.34, P = 0.002; 10-fold cross-validation: R = 0.29, P = 0.009; split-half design: R = 0.31, P = 0.026, with two subcohorts matched by baseline UPDRS-III scores, age and DBS improvements, see Methods). Figure 3c contrasts two example coupling profiles from a top and a poor responding patient, respectively. Given that hemispheres were treated as individual data points in these analyses and to account for within-subject associations, we also computed a leave-one-patient out cross-validation, which yielded a similar result (R = 0.31, P = 0.003).

When comparing the high beta average map to the R-map, grand average high beta coupling peaked in the motor cortex, while motor cortex coupling seemed to be less beneficial for clinical improvement in the R-map (Figure 2d; Figure 3a). To gain a deeper understanding of this discrepancy, we mapped the variance of coupling intensities (across hemispheres) onto the surface (supplementary figure S2). The variance peaked in the mesial motor cortex, exactly where we found strong average coupling. This indicates, that while the average coupling was dominated by motor cortex high beta, it was also the location with greatest variance. Together, these findings indicate that i) coupling to motor cortex is high on average but ii) shows high variance and iii) is low in patients with optimal improvements. Similarly, the mesial prefrontal peak-location of the R-map (Figure 3a) showed a peak in variance. To further investigate the relevance of each of these two variance peak regions for clinical outcomes, we constrained the correlation of amplitude coupling and DBS improvements to these two regions of interest. This analysis revealed positive and significant relationships between amplitude coupling in the mesial prefrontal region and DBS improvement (R = 0.23; P = 0.024), but no significant relationship for mesial motor cortex coupling (R = −0.10; P = 0.310).

To examine whether the results were driven by specific symptoms captured in the UPDRS-III, we repeated the analysis correlating individual amplitude coupling maps, with symptom-specific sub-scores for bradykinesia, rigidity and tremor, respectively. In agreement with the consistent association of akinetic-rigid symptoms with local and network beta synchrony^17,26,27^, the bradykinesia and rigidity R-maps showed a similar overall pattern as the total UPDRS R-map (supplementary figure S3). Leave-one-out cross-validations of rigidity and bradykinesia R-maps were significant (rigidity: R = 0.41; P = 0.003; bradykinesia: R = 0.26; P = 0.030), while the tremor R-map did not cross-validate (R = 0.12; P = 1.00).

Additionally, we tested whether the approach could estimate DBS outcomes across DBS centers, surgeons and even different MEG devices, i.e. would withstand potential systematic biases that may have been introduced by pooling of subcohorts from different centers. We first calculated the model on the larger cohort with n = 80 hemispheres from Düsseldorf to then estimate outcomes in the n = 20 hemispheres from Berlin (Figure 4). In this analysis, cross-estimates significantly correlated with empirical improvements, as well (R = 0.74; P_(1)_ = 8.9e-5). For sake of completeness, we also built a model based on the small cohort (n = 20) and attempted to predict variance in the larger cohort (n = 80). As expected based on prior work^9^, we predicted less variance, but still found a significant association (R = 0.20; P_(1)_ = 0.035; supplementary figure S4).

**Figure 4.**
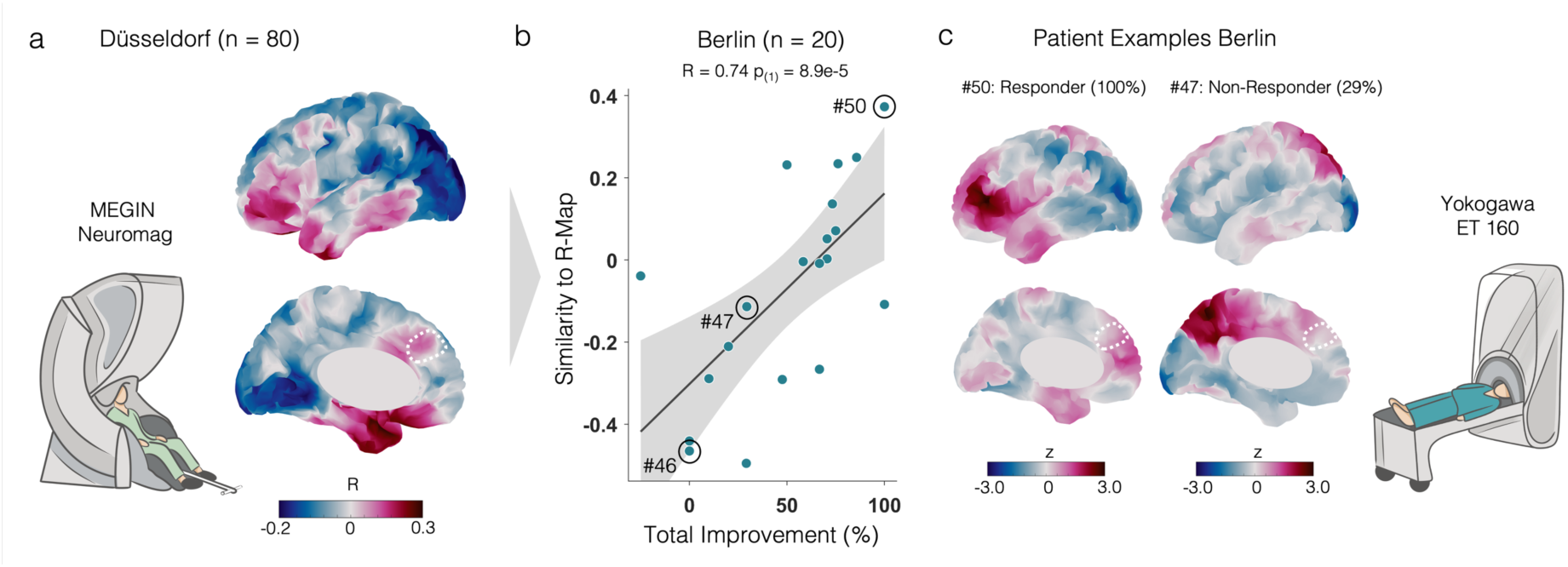
Electrophysiological DBS network map estimates outcomes across DBS centers, surgeons and MEG devices. **a** The R-map was generated based on the larger patient cohort from Düsseldorf (recordings in 80 hemispheres). Individual coupling maps from the Berlin cohort (n = 20 hemispheres) were spatially correlated with the Düsseldorf R-map. **b** Similarity of patient-specific coupling maps in the Berlin cohort to the R-map generated from the Düsseldorf cohort demonstrated a significant correlation with the empirical stimulation improvements. Grey shaded areas represent 95% confidence intervals. **c** Two example coupling maps illustrate patients with optimal and suboptimal stimulation responses. Dashed lines mark the same area that was highlighted in Figure 3, indicating the prefrontal peak region to which an electrode should ideally be connected to achieve maximal treatment benefit for a patient. The coupling map for patient # 46 marked in the plot is provided in figure 3c. **a,c** Drawings of the two different MEG systems that were used to acquire the data at the two centers are depicted next to the respective cohort. Data in Düsseldorf was acquired in the upright position using a Neuromag system from MEGIN and patients in Berlin were recorded in the supine position using a Yokogawa MEG system.

### Electrophysiological and MRI-based DBS network maps align spatially

As mentioned in the introduction, the spatial topographies of the Parkinson’s disease response network have been extensively mapped using fMRI. However, given limitations of this method, its temporal characteristics (i.e. the predominant frequency band in which it operates) had not been mapped. Hence, it was a key next step to align our electrophysiologically defined response network with the published fMRI networks. Indeed, the high beta R-map identified here spatially resembled published MRI-based DBS R-maps^8,10^. To quantify the alignment between the optimal cortical STN connectivity pattern derived from amplitude coupling and fMRI, we spatially correlated electrophysiological and MRI-based maps in a vertex-wise manner which showed clear spatial similarity (spatial correlation fMRI R-map vs. neurophysiology R-map: R = 0.40). This relationship was more similar than could be expected by chance (following the permutation-based test introduced in^47^; P = 0.039), and was substantially greater than similarities between theta-alpha-, low beta- and the fMRI based R-map respectively (spatial correlation theta-alpha R = −0.31; low-beta R = −0.06). Importantly, the fMRI R-map was calculated on an independent patient cohort not included in the MEG analysis for this study^8^. The two main components that aligned particularly well across both maps comprised the negative correlations within the motor cortex, and the positive correlations in prefrontal areas (Figure 5a-c).

**Figure 5.**
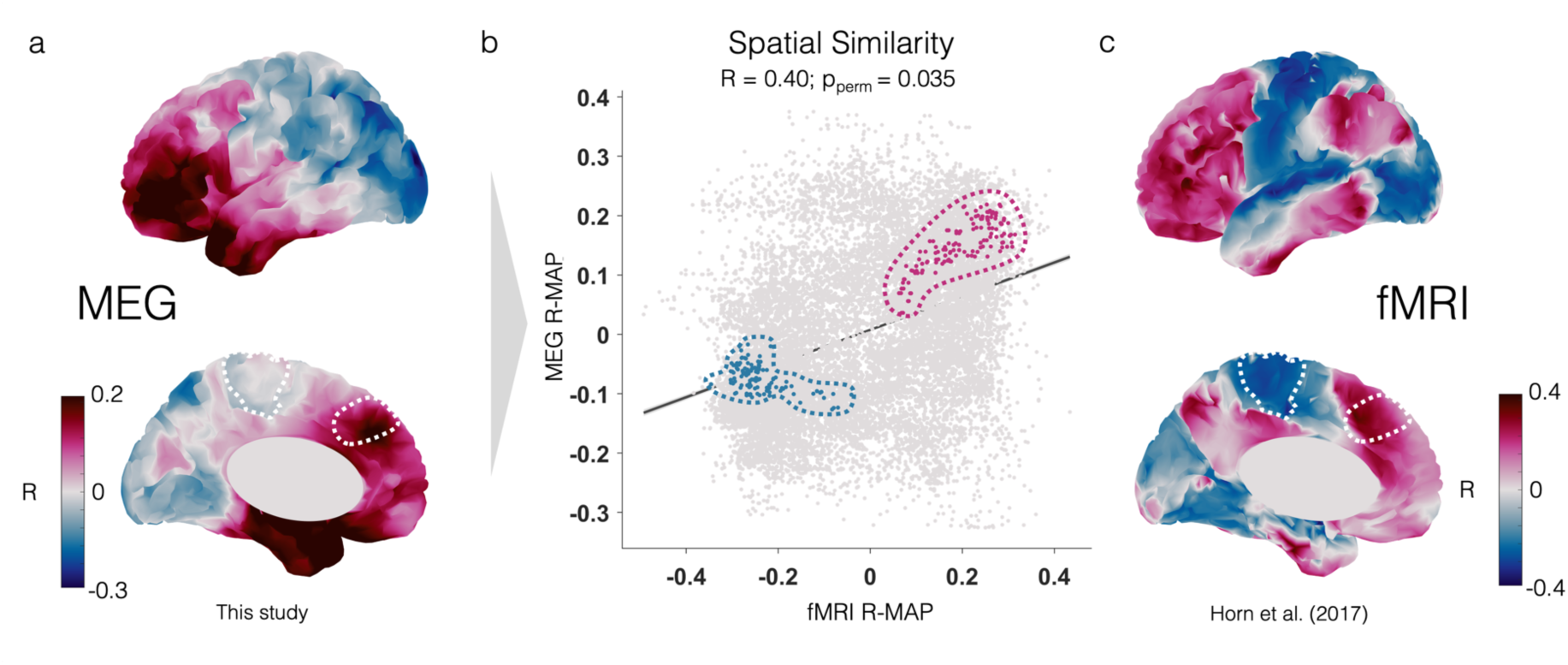
Spatial correlation between MEG and fMRI R-maps. **a,c** Magnetoencephalography (MEG) and functional magnetic resonance imaging (fMRI) derived correlation maps (R-maps) based on stimulation improvements. The fMRI based R-map was previously published with another patient cohort not included in the MEG analysis for this study^8^. **b** Spatial correlation between the cortical vertices in both, the MEG-derived and the fMRI R-maps and the permutation-based P-value (P_perm_) are reported. Blue colored dots surrounded by the dashed line indicate mesial motor vertices in both maps, while red colored dots indicate vertices in the prefrontal mesial cortex. Both areas are negatively (blue) and positively (red) correlated in both R-maps.

## Discussion

This work builds upon unique multi-center human data of combined invasive STN and non-invasive MEG recordings (N = 100 hemispheres) with the goal to define stimulation response in subthalamic DBS for Parkinson’s disease with high precision in both *spatial* and *temporal* domains. Three main conclusions can be drawn from this study. First, subthalamic stimulation sites that were coupled to a distributed brain network in the high beta frequency band led to better clinical outcomes after DBS. Second, this relationship was robust across DBS centers, surgeons and different MEG devices. Third, the topography of this high beta-band network matched the topography of a previously derived fMRI-based network, linking prior DBS studies that had focused on either electrophysiology or neuroimaging alone. Taken together, these results provide cross-modal insight into the DBS response of Parkinson’s disease, showing that effective stimulation sites are maximally engaged with a specific brain network in the high beta frequency band.

The electrophysiological DBS network map provides the following key insights: First, we were able to broadly replicate the fMRI-based DBS response network using combined LFP-MEG recordings. Second, the refined definition of the electrophysiological network puts stronger focus on frontal regions while deemphasizing potentially spuriously correlated parietal cortical regions that were found using fMRI. Third, and most importantly, the network identified here is based on a time scale *two orders of magnitude* faster than the fMRI network described previously. As outlined above, it might be expected that, in electrophysiological terms, this network would operate in the high beta frequency, but this had not been empirically demonstrated before. Moreover, the identified network might be useful to guide DBS programming and maybe even DBS targeting, in the future. For DBS programming, novel sensing-enabled DBS devices allow LFP recordings from each stimulation contact without externalization^26,28^. These data could be combined with EEG or even MEG^55^ to evaluate contact-specific connectivity patterns and guide contact selection. For DBS targeting, an intraoperative setup combining frontal EEG or electrocorticography (ECoG) with subthalamic LFP recordings could be conceived with the aim of monitoring connectivity while implanting electrodes along stereotactic trajectories.

Synchronous beta band activity has emerged as a hallmark pathophysiological network signature that characterizes and correlates with akinetic-rigid symptomatology in Parkinson’s disease ^25,56^. As summarized by Binns and colleagues, subthalamic *low* beta power is typically attenuated by the intake of dopaminergic medication, while DBS leads to an attenuation of *high* beta synchrony^21^. High beta coherence has been described to dominate STN-cortical synchrony in the context of Parkinson’s disease ^13,15,16^. Localized low beta *power*, in turn, has been attributed to symptom severity, especially in the context of bradykinesia, when recorded from the STN^27^. Several other electrophysiological studies that combined subcortical and cortical recordings have observed changes in cortico-STN coherence due to DBS and dopaminergic medication^13,21,57^. However, thus far, few studies have related these signatures to clinical baseline scores or DBS improvements^13,17,58^. One study reported negative correlations between motor cortex-STN beta coherence and akinetic-rigid symptoms, while another identified high frequency as well as low gamma cortex-STN coherence as predictive features of the DBS outcome^17,58^. The lack of robust associations between clinical scores and (beta) coherence metrics suggests that most of the changes in coherence observed in prior studies represent dynamics that were neither causally related to symptomatology nor to the treatment effect. Instead, the observed effects could have been compensatory in nature, or spuriously correlated with the state of DBS, without exerting impact on clinical symptoms^59^. Instead of relying on absolute *magnitudes* of coupling with individual cortical regions to predict DBS effects, the MRI-based DBS network mapping field has leveraged the *spatial pattern* across the entire cortex (or brain) to employ *topologies* rather than *absolute values*^8,10,60,61^.

Our study applies this same concept to electrophysiological measures of connectivity between the STN and cortex, and by doing so, is focused on characterizing an optimal DBS response network in both temporal frequency and anatomical space. Indeed, we identified a specific network that was coupled to optimal subthalamic stimulation sites in the high beta range and demonstrated remarkably robust utility to estimate variance in clinical outcomes in unseen patients. Similar to published fMRI-based DBS network maps^8–10^, the network featured strong coupling to mesial prefrontal and other frontal areas. In agreement with the MRI-based literature^8–10^, strong coupling to the primary motor cortex was negatively correlated with DBS outcomes (Figure 3a) and the map we identified here using electrophysiology was more similar to the published fMRI based map than could be expected by chance (Figure 5).

DBS response networks based on fMRI have been proposed to inform noninvasive neuromodulation targets^62^. To test this hypothesis in Parkinson’s disease, a recent prospective randomized double-blinded trial noninvasively stimulated the fMRI defined *DBS response network* which indeed led to significantly higher motor symptom improvements compared to sham stimulation^24^. The same strategy to define noninvasive treatment networks based on invasive DBS data has been successful in other indications, such as major depression, where it even led to novel FDA approvals^63–65^. Besides the similar *spatial* pattern observed in our data, the DBS response network we identify now shows *frequency-specificity* for the high beta band. This could extend the available information to more deliberately guide non-invasive stimulation paradigms, by e.g. targeting high beta temporal dynamics above and beyond the anatomically defined multifocal stimulation target^24^. For instance, a refined multifocal transcranial alternating current stimulation (tACS) protocol could be applied in the high beta range on a *temporal* scale, while *spatially* targeting the DBS response network. Alternatively, or additionally, such paradigms could be applied in a closed-loop fashion using a steering signal recorded in the high beta range to for instance apply anti-phasic tACS, or apply transcranial magnetic stimulation in a phase-specific manner.

Several limitations should be considered when interpreting our results. First, our study followed a retrospective design and patients with missing follow-up visits and clinical data needed to be excluded from the analysis. Further, as this study is the first to describe the electrophysiological DBS response network, we were not able to assess its utility in a prospective manner. Future studies are needed to prospectively validate the clinical translatability of our model for DBS programming or surgical targeting, optimally in randomized controlled studies. Second, this study setup required access to specialized equipment that does not scale well for clinical application. These combined MEG-LFP recordings have only been acquired at few centers world-wide. This highlights the need for future replication with simpler and/or more accessible equipment such as combined LFP-EEG recordings using sensing enabled neurostimulators. Third, even though we find similarities between our R-maps and the a priori published fMRI R-map, our findings cannot definitely resolve whether the patterns we observed in the MEG-derived maps truly represent the same neurophysiological underpinnings across modalities. This being said, a recent within-subject study on resting-state networks in MEG and fMRI demonstrated high correspondence between cortico-cortical amplitude coupling and fMRI resting state networks^34^.

In summary, we present a novel approach to apply DBS network mapping using electrophysiological data from a unique dataset of invasive human STN LFP recordings acquired in conjunction with MEG. We identify a previously unknown pathophysiological network target for subthalamic DBS in Parkinson’s disease that operates in the high beta band and relates to DBS symptom improvement. Future studies might benefit from the proposed mapping approach and identified electrophysiological network profile, enabling more precise and potentially non-invasive targeting of dysfunctional Parkinsonian circuits in both the *spatial* and *temporal* domain.

## Data Availability

The patient imaging and electrophysiology data cannot be publicly shared, since this would compromise patient privacy according to current data protection regulations. They are, however, available from the principal investigators of the collecting sites upon reasonable request within the framework of a data-sharing agreement and the respective local ethical regulations. Inquiries for further information and data-sharing requests should be directed to the corresponding authors of this manuscript who commit to replying to any request within a timeframe of 30 d.

## Acknowledgements

BHB, LLG and BH gratefully acknowledge support by the Prof. Dr. Klaus Thiemann Foundation (Parkinson Fellowship 2022, 2023 and 2024). RL was supported by the Deutsche Forschungsgemeinschaft (DFG, German Research Foundation) – Project-ID 424778381 – TRR 295 and the Berlin Institute of Health. EF gratefully acknowledges support from the Volkswagen Foundation (Lichtenberg program 89387) and the Deutsche Forschungsgemeinschaft (DFG, German Research Foundation) – Project-ID 424778381 – TRR 295. KJM was supported by National Institutes of Health (NIH, U01-NS128612) and the Foundation for OCD Research (FFOR). AH was supported by the German Research Foundation (Deutsche Forschungsgemeinschaft, 424778381 – TRR 295), Deutsches Zentrum für Luft-und Raumfahrt (DynaSti grant within the EU Joint Programme Neurodegenerative Disease Research, JPND), the NIH (R01MH130666, 1R01NS127892-01, 2R01 MH113929 & UM1NS132358) as well as the New Venture Fund (FFOR Seed Grant). Manuscript contents are solely the responsibility of the authors and do not necessarily represent the official views of the funders, who had no role in study design, data collection and analysis, decision to publish, or preparation of the manuscript. We would like to thank Dora Hermes for insightful methodological discussions.

## Supplementary material

**Table S1.** Patient Characteristics.

| Patient | DBS Center | UPDRS Stim OFF | UPDRS Stim ON | Contact R | Contact L | Amp R | Amp L | Pulse Width | Freq | Electrode model |
| --- | --- | --- | --- | --- | --- | --- | --- | --- | --- | --- |
| 1 | DUS | 42 | 18 | 10B | 10A | 2.1 mA | 1.9 mA | 60 | 130 | Abbott 6172 |
| 2 | DUS | 23 | 17 | 10C | 2B | 1.4 mA | 1.2 mA | 60 | 130 | Abbott 6172 |
| 3 | DUS | 34 | 26 | 10A | 1 | 1.3 mA | 1.3 mA | 60 | 130 | Abbott 6172 |
| 4 | DUS | 34 | 8 | 10ABC | 2ABC | 1.7 mA | 2.8 mA | 60 | 130 | Abbott 6172 |
| 5 | DUS | 53 | 34 | 10A | 2A | 3.5 mA | 4.0 mA | 40 | 180 | Abbott 6172 |
| 6 | DUS | 30 | 26 | 2C | 10B | 1.5 mA | 1.5 mA | 30 | 130 | Abbott 6172 |
| 7 | DUS | 53 | 26 | 10A | 2A | 2.2 mA | 3.2 mA | 40 | 130 | Abbott 6172 |
| 8 | DUS | 48 | 33 | 11A | 3B | 1.25 mA | 0.75 mA | 60 | 130 | Abbott 6172 |
| 9 | DUS | 39 | 29 | 10AC | 2AC | 2.95 mA | 2.2 mA | 60 | 125 | Abbott 6172 |
| 10 | DUS | 50 | 31 | 10ABC | 2ABC | 3.0 mA | 3.0 mA | 50 | 130 | Abbott 6172 |
| 11 | DUS | 29 | 26 | 10B | 1 | 2.4 mA | 3.6 mA | 60 | 60 | Abbott 6172 |
| 12 | DUS | 55 | 30 | 10A | 2A | 1.4 mA | 2.5 mA | 60 | 130 | Abbott 6172 |
| 13 | DUS | 42 | 31 | 10ABC | 2C | 3.0 mA | 2.2 mA | 60 | 130 | Abbott 6172 |
| 14 | DUS | 28 | 18 | 10ABC | 3ABC | 3.0 mA | 3.3 mA | 60 | 130 | Abbott 6172 |
| 15 | DUS | 47 | 28 | 12, 11C | 4, 3B | 3.8 mA | 3.9 mA | 60 | 116 | Abbott 6172 |
| 16 | DUS | 40 | 30 | 11B | 3ABC | 2.1 mA | 1.8 mA | 60 | 130 | Abbott 6172 |
| 17 | DUS | 13 | 11 | 10AC | 1 | 1.3 mA | 1.8 mA | 60 | 130 | Abbott 6172 |
| 18 | DUS | 48 | 45 | 10ABC | 1 | 0.8 mA | 1.5 mA | 60 | 130 | Abbott 6172 |
| 19 | DUS | 44 | 45 | 11C | 3ABC | 2.4 mA | 3.6 mA | 60 | 130 | Abbott 6172 |
| 20 | DUS | 34 | 16 | 2B | 11ABC | 1.2 mA | 4.5 mA | 40 | 130 | Abbott 6172 |
| 21 | DUS | 49 | 32 | 1 | 2 | 3.7 V | 3.7 V | 60 | 130 | Medtronic 3389 |
| 22 | DUS | 23 | 15 | 1 | 1 | 2.1 V | 2.4 V | 60 | 130 | Medtronic 3389 |
| 23 | DUS | 34 | 21 | 1,2 | 1,2 | 2.9V | 3.0V | 60 | 130 | Medtronic 3389 |
| 24 | DUS | 46 | 37 | 2,3 | 2,3 | 3.9V | 3.4 V | 60 | 130 | Medtronic 3389 |
| 25 | DUS | 25 | 13 | 3 | 2 | 2.0V | 2.3V | 60 | 130 | Medtronic 3389 |
| 26 | DUS | 51 | 29 | 2,3 | 2 | 3.2V | 2.5 V | 60 | 130 | Medtronic 3389 |
| 27 | DUS | 34 | 22 | 3 | 2 | 2.0V | 2.0 V | 60 | 130 | Medtronic 3389 |
| 28 | DUS | 33 | 12 | 2 | 1 | 2.9 V | 2.9 V | 90 | 140 | Medtronic 3389 |
| 29 | DUS | 38 | 19 | 1 | 1 | 2.9 V | 2.3 V | 60 | 130 | Medtronic 3389 |
| 30 | DUS | 22 | 10 | 2 | 2 | 1.3 | 0.7 | 62 | 130 | St. Jude Active Tip |
| 31 | DUS | 44 | 28 | 1 | 1 | 2.4 V | 2.1 V | 60 | 130 | Medtronic 3389 |
| 32 | DUS | 50 | 34 | 4 | 4 | 3.1 | 2.7 | 60 | 130 | Boston Scientific |
| 33 | DUS | 35 | 12 | 3 | 3 | 0.5 | 3.5 | 60 | 130 | Boston Scientific |
| 34 | DUS | 57 | 40 | 2 | 3 | 3.1 V | 3.3 V | 60 | 130 | Medtronic 3389 |
| 35 | DUS | 34 | 27 | 2 | 2 | 2.0 | 2.0 | 90 | 130 | St. Jude Active Tip |
| 36 | DUS | 50 | 31 | 1 | 1 | 2.1V | 2.5 V | 60 | 130 | Medtronic 3389 |
| 37 | DUS | 57 | 17 | 0 | 0 | 1.4 V | 3.8 V | 60 | 130 | Medtronic 3389 |
| 38 | DUS | 38 | 15 | 1 | 1 | 2.1 V | 2.5 V | 60 | 130 | Medtronic 3389 |
| 39 | DUS | 58 | 36 | 1,3 | 0,1 | 1.9/2.4 mA | 2.5/1.5mA | 60 | 125 | Medtronic 3389 |
| 40 | DUS | 40 | 18 | 1 | 1 | 2.5 V | 1.8 V | 60 | 130 | Medtronic 3389 |
| 41 | BER | 34 | 16 | 1 | 4,5 | 3.58 V | 2.35 V | 60 | 130 | Medtronic 3389 |
| 42 | BER | 103 | 32 | 2 | 10 | 2.9 V | 2.6 V | 60 | 130 | Medtronic 3389 |
| 43 | BER | 36 | 11 | 0,1 | 8,9 | 2.0 V | 2.0 V | 60 | 130 | Medtronic 3389 |
| 44 | BER | 63 | 49 | 3 | 11 | 1.8 V | 2.0 V | 60 | 130 | Medtronic 3389 |
| 45 | BER | 18 | 15 | 0 | 8 | 2.2 V | 2.5 V | 60 | 90 | Medtronic 3389 |
| 46 | BER | 37 | 24 | 1 | 9 | 3.8 V | 3.0 V | 60 | 130 | Medtronic 3389 |
| 47 | BER | 61 | 46 | 2 | 10 | 2.2 V | 2.0 V | 60 | 130 | Medtronic 3389 |
| 48 | BER | 38 | 8 | 12 | 4 | 2.2 V | 3.8 V | 60 | 130 | Medtronic 3389 |
| 49 | BER | 72 | 22 | 13,14,15 | 5,6,7 | 3.0 mA | 2.2 mA | 60 | 130 | Boston 2202 |
| 50 | BER | 20 | 1 | 3 | 11 | 3.3 mA | 3.2 mA | 60 | 130 | Boston 2202 |
| **Mean** |  | **41,7** | **24,4** |  |  |  |  |  |  |  |
| **Std** |  | **15,3** | **11** |  |  |  |  |  |  |  |

**Table S2.**
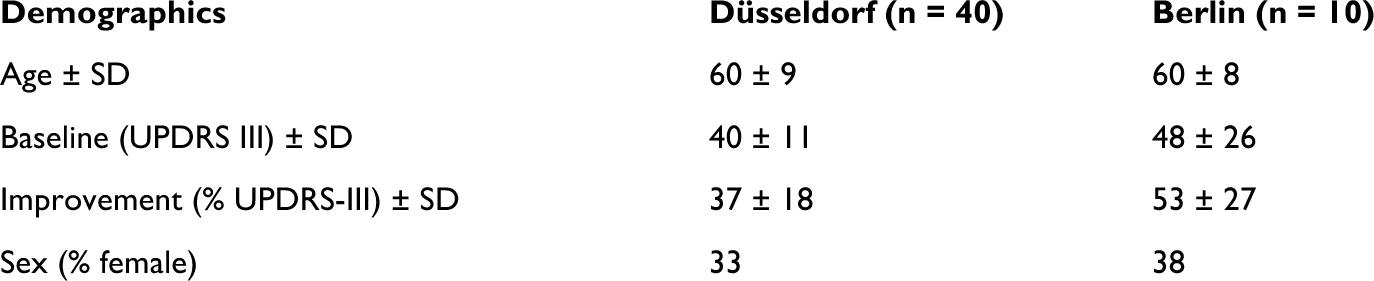
Cohort demographics.

| Demographics | Düsseldorf (n = 40) | Berlin (n = 10) |
| --- | --- | --- |
| Age ± SD | 60 ± 9 | 60 ± 8 |
| Baseline (UPDRS III) ± SD | 40 ± 11 | 48 ± 26 |
| Improvement (% UPDRS-III) ± SD | 37 ± 18 | 53 ± 27 |
| Sex (% female) | 33 | 38 |

**Figure S1.**
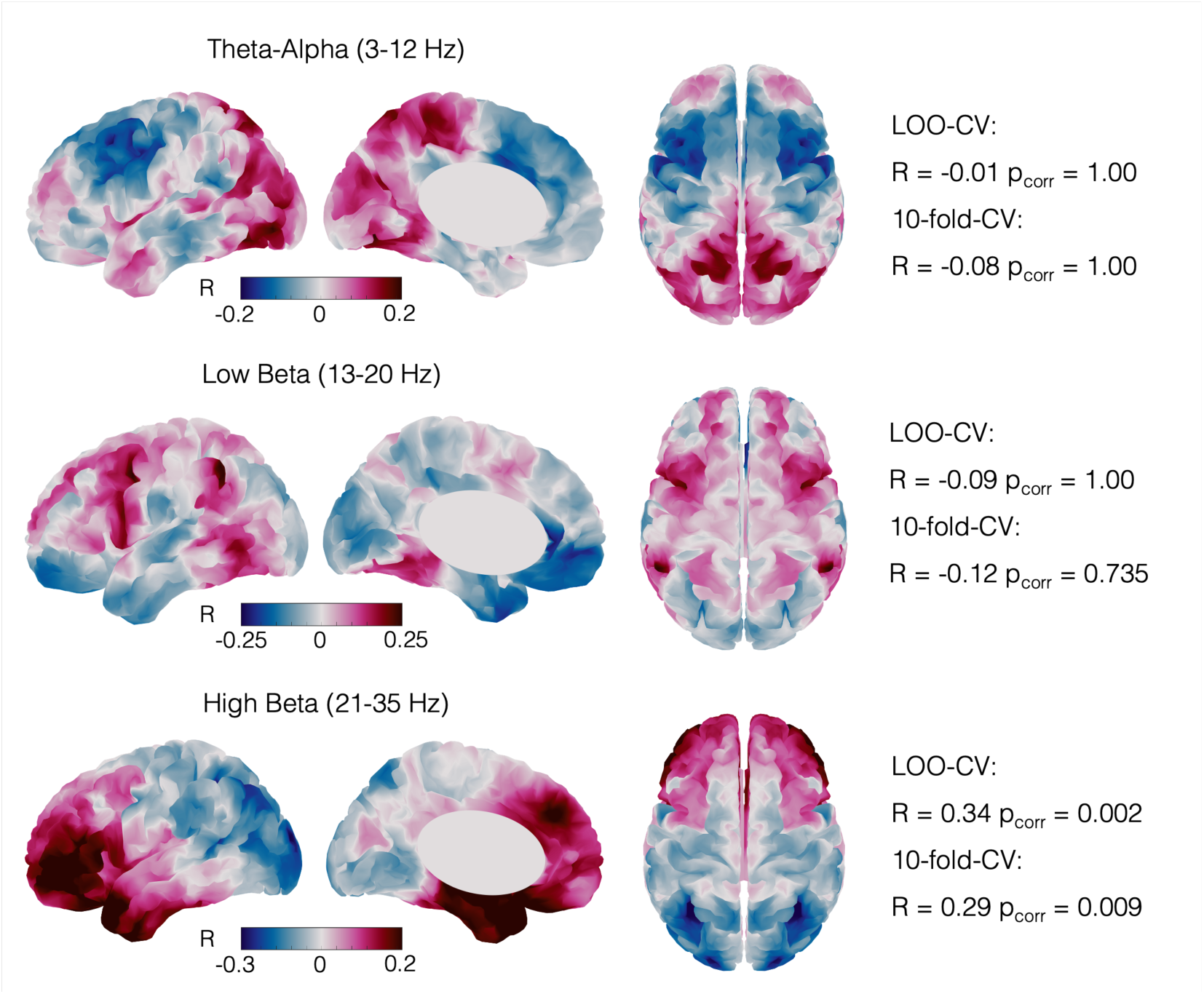
R-maps across frequency bands. Correlation maps (R-maps) for each of the three canonical frequency bands. Leave-one-out-cross-validations (LOO-CV) and 10-fold-cross-validation results are reported alongside the figures and P-values are Bonferroni-corrected for multiple comparisons.

**Figure S2.**
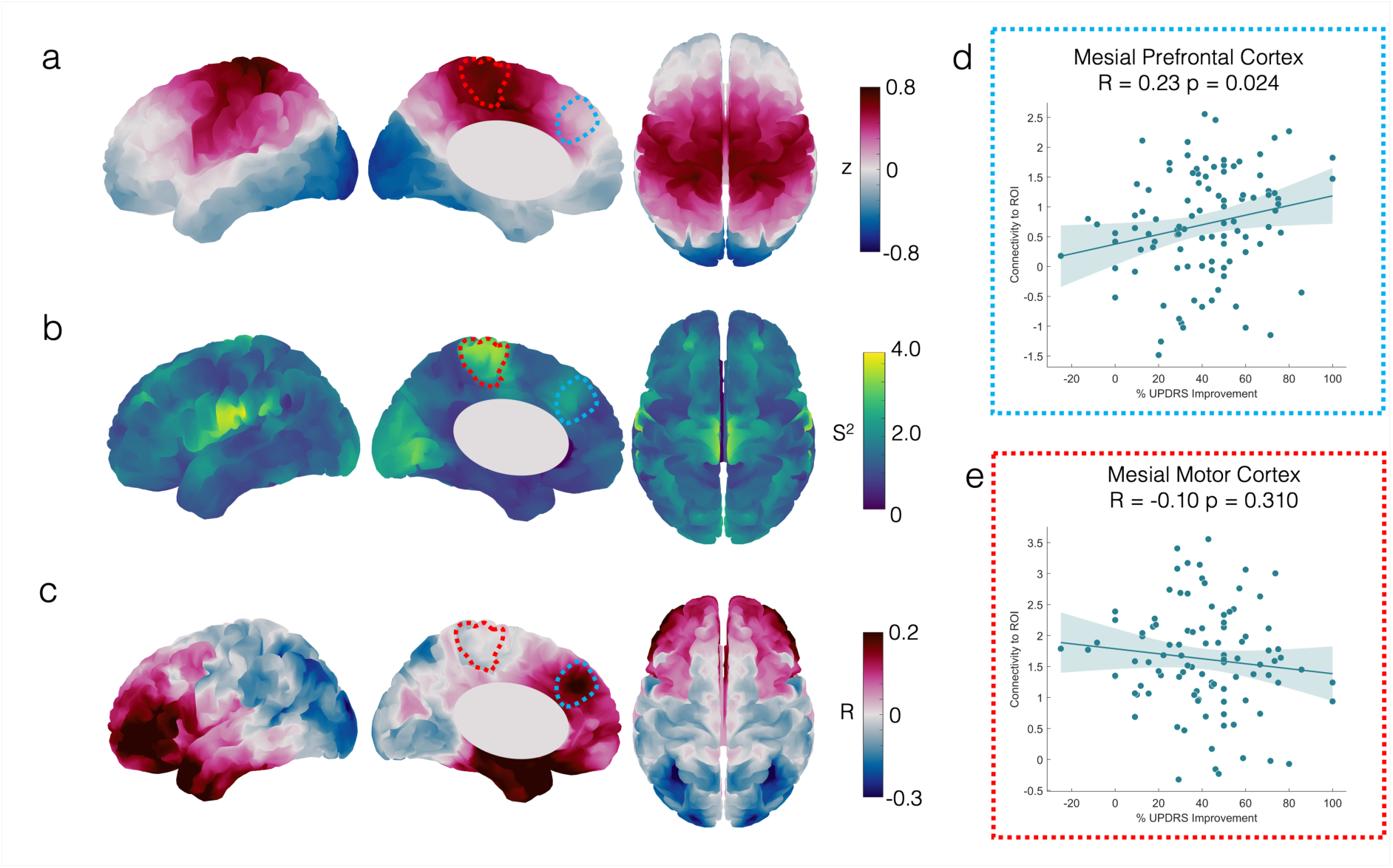
Average, Variance and R-map. a Average coupling map in the high beta band across 100 hemispheres, b Variance map in the high beta band, showing variance of high beta coupling across 100 hemispheres, c correlation map (R-map) for coupling in the high beta band correlated with DBS improvements. d region of interest (ROI) maximal high beta coupling in mesial prefrontal cortex (see white dotted line in a-c) across hemispheres correlates with DBS hemiscore improvements, e ROI maximal high beta coupling in mesial motor cortex does not correlate with DBS improvements.

**Figure S3.**
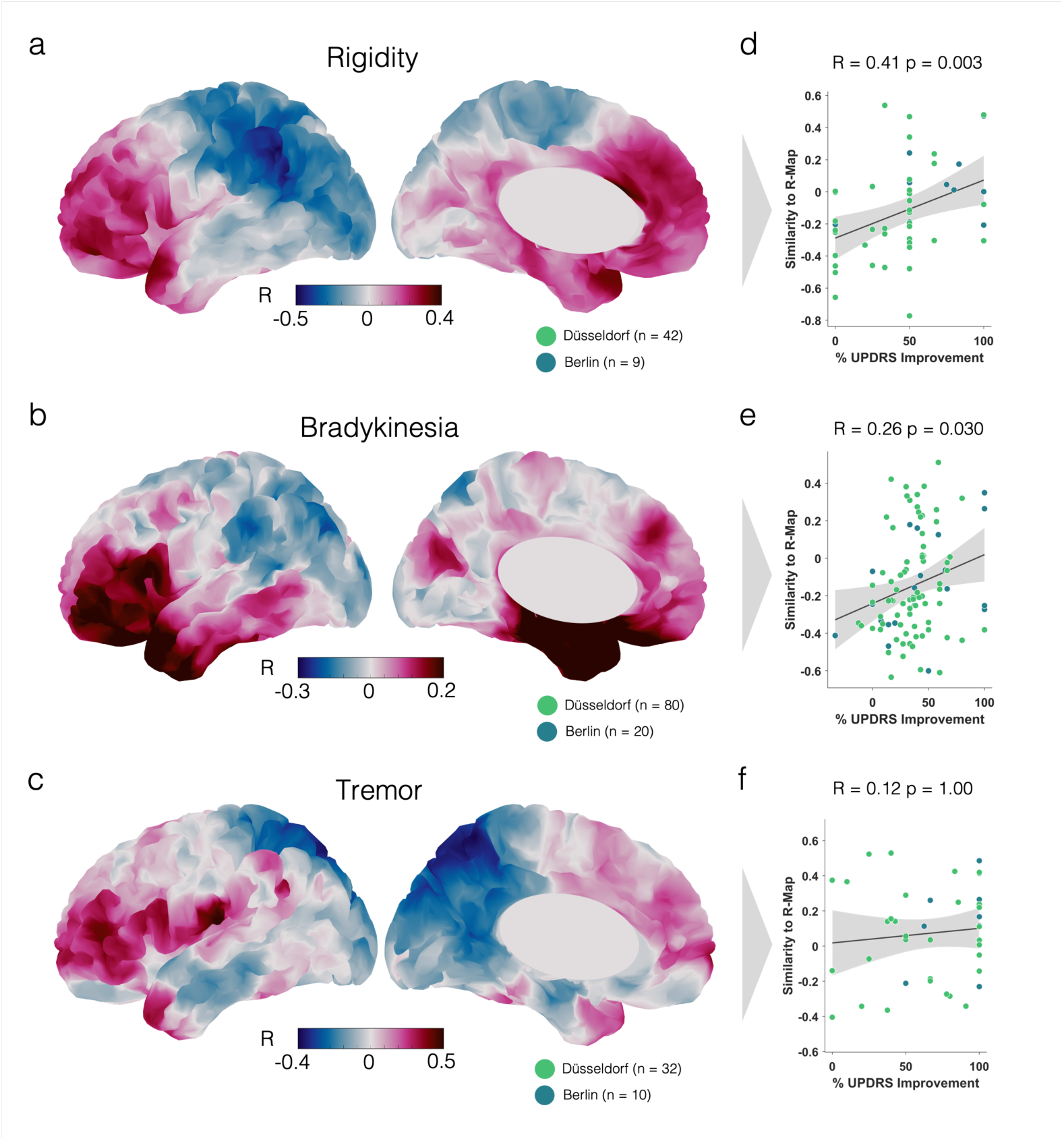
Symptom-specific R-maps. a-c. R-maps based on symptom-sub-score improvements in rigidity (a), bradykinesia (b) and tremor (c). The number of included hemispheres depended on the presence of baseline symptoms. That resulted in smaller cohorts-especially for tremor and rigidity. **d-f** Leave-one-out cross-validations for the three symptom-specific R-maps. P-values are Bonferroni-corrected.

**Figure S4.**
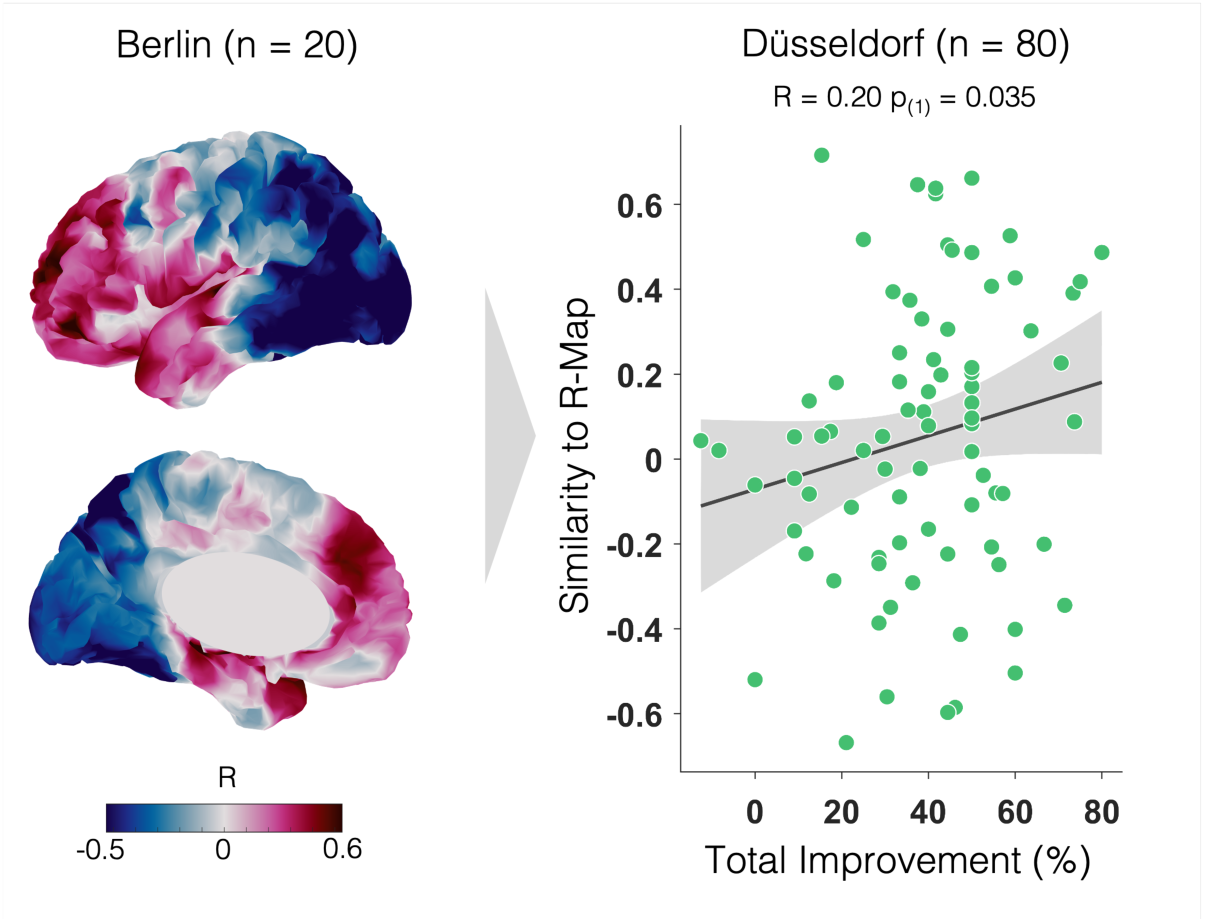
Cross-prediction Berlin R-map. The R-map based on the Berlin cohort (n =20 hemispheres) was spatially correlated with the fingerprints from the Düsseldorf cohort (n = 80 hemispheres) to estimate empirical UPDRS-III improvements. The correlation between estimated (Similarity to R-map) and empirical improvements (total improvement) only showed a positive trend. As discussed in the main text, models with less than 30 data points are normally not robust enough to predict unseen, external data.

## References

1. Deuschl G, Schade-Brittinger C, Krack P, et al. A randomized trial of deep-brain stimulation for Parkinson’s disease. The New England Journal of Medicine. 2006;355(9):896–908. doi:10.1056/NEJMoa060281

2. Tödt I, Al-Fatly B, Granert O, et al. The Contribution of Subthalamic Nucleus Deep Brain Stimulation to the Improvement in Motor Functions and Quality of Life. Movement Disorders. 2022;37(2):291–301. doi:10.1002/MDS.28952

3. Treu S, Strange B, Oxenford S, et al. Deep brain stimulation: Imaging on a group level. NeuroImage. 2020;219:117018. doi:10.1016/j.neuroimage.2020.117018

4. Roediger J, Dembek TA, Wenzel G, Butenko K, Kühn AA, Horn A. STIMFIT— A Data-Driven Algorithm for Automated Deep Brain Stimulation Programming. Movement Disorders. 2022;37(3):574–584. doi:10.1002/mds.28878

5. Benabid AL, Pollak P, Louveau A, Henry S, de Rougemont J. Combined (thalamotomy and stimulation) stereotactic surgery of the VIM thalamic nucleus for bilateral Parkinson disease. Appl Neurophysiol. 1987;50(1-6):344–346.

6. Hassler R, Riechert T, Mundinger F, Umbach W, Ganglberger JA. PHYSIOLOGICAL OBSERVATIONS IN STEREOTAXIC OPERATIONS IN EXTRAPYRAMIDAL MOTOR DISTURBANCES. Brain. 1960;83(2):337–350. doi:10.1093/brain/83.2.337

7. Henderson JM. “Connectomic surgery”: diffusion tensor imaging (DTI) tractography as a targeting modality for surgical modulation of neural networks. Front Integr Neurosci. 2012;6. doi:10.3389/fnint.2012.00015

8. Horn A, Reich M, Vorwerk J, et al. Connectivity Predicts deep brain stimulation outcome in Parkinson disease: DBS Outcome in PD. Ann Neurol. 2017;82(1):67–78. doi:10.1002/ana.24974

9. Wang Q, Akram H, Muthuraman M, et al. Normative vs. patient-specific brain connectivity in deep brain stimulation. NeuroImage. 2021;224:117307. doi:10.1016/j.neuroimage.2020.117307

10. Sobesky L, Goede L, Odekerken VJJ, et al. Subthalamic and pallidal deep brain stimulation: are we modulating the same network? Brain. 2021;145(1):251–262. doi:10.1093/BRAIN/AWAB258

11. Rajamani N, Friedrich H, Butenko K, et al. Deep brain stimulation of symptom-specific networks in Parkinson’s disease. Nat Commun. 2024;15(1):4662. doi:10.1038/s41467-024-48731-1

12. Hollunder B, Ostrem JL, Sahin IA, et al. Mapping dysfunctional circuits in the frontal cortex using deep brain stimulation. Nature Neuroscience. 2024;27(3):573–586. doi:10.1038/s41593-024-01570-1

13. Oswal A, Beudel M, Zrinzo L, et al. Deep brain stimulation modulates synchrony within spatially and spectrally distinct resting state networks in Parkinson’s disease. Brain. 2016;139(5):1482–1496. doi:10.1093/BRAIN/AWW048

14. Oswal A, Cao C, Yeh CH, et al. Neural signatures of hyperdirect pathway activity in Parkinson’s disease. Nature Communications. 2021;12(1):1–14. doi:10.1038/s41467-021-25366-0

15. Litvak V, Jha A, Eusebio A, et al. Resting oscillatory cortico-subthalamic connectivity in patients with Parkinson’s disease. Brain. 2011;134(2):359–374. doi:10.1093/BRAIN/AWQ332

16. Hirschmann J, Özkurt TE, Butz M, et al. Distinct oscillatory STN-cortical loops revealed by simultaneous MEG and local field potential recordings in patients with Parkinson’s disease. Neuroimage. 2011;55(3):1159–1168. doi:10.1016/j.neuroimage.2010.11.063

17. Hirschmann J, Özkurt TE, Butz M, et al. Differential modulation of STN-cortical and cortico-muscular coherence by movement and levodopa in Parkinson’s disease. NeuroImage. 2013;68:203–213. doi:10.1016/J.NEUROIMAGE.2012.11.036

18. Sharma A, Vidaurre D, Vesper J, Schnitzler A, Florin E. Differential dopaminergic modulation of spontaneous cortico–subthalamic activity in parkinson’s disease. eLife. 2021;10. doi:10.7554/ELIFE.66057

19. van Wijk BCM, Neumann WJ, Kroneberg D, et al. Functional connectivity maps of theta/alpha and beta coherence within the subthalamic nucleus region. NeuroImage. 2022;257:119320. doi:10.1016/J.NEUROIMAGE.2022.119320

20. Cao C, Litvak V, Zhan S, et al. Low-beta versus high-beta band cortico-subcortical coherence in movement inhibition and expectation. Neurobiology of Disease. 2024;201:106689. doi:10.1016/j.nbd.2024.106689

21. Binns TS, Köhler RM, Vanhoecke J, et al. Shared pathway-specific network mechanisms of dopamine and deep brain stimulation for the treatment of Parkinson’s disease. bioRxiv. Published online April 2024:2024.04.14.586969. doi:10.1101/2024.04.14.586969

22. Horn A, Fox MD. Opportunities of connectomic neuromodulation. NeuroImage. 2020;221:117180. doi:10.1016/j.neuroimage.2020.117180

23. Hollunder B, Rajamani N, Siddiqi SH, et al. Toward personalized medicine in connectomic deep brain stimulation. Progress in Neurobiology. 2022;210:102211. doi:10.1016/J.PNEUROBIO.2021.102211

24. Goede LL, Oxenford S, Kroneberg D, et al. Linking Invasive and Noninvasive Brain Stimulation in Parkinson’s Disease: A Randomized Trial. Movement Disorders. Published online July 2024. doi:10.1002/MDS.29940

25. Brown P. Oscillatory nature of human basal ganglia activity: Relationship to the pathophysiology of Parkinson’s disease. Movement Disorders. 2003;18(4):357–363. doi:10.1002/MDS.10358

26. Feldmann LK, Lofredi R, Neumann WJJ, et al. Toward therapeutic electrophysiology: beta-band suppression as a biomarker in chronic local field potential recordings. npj Parkinson’s Disease. 2022;8(1):1–9. doi:10.1038/s41531-022-00301-2

27. Kühn AA, Kupsch A, Schneider GH, Brown P. Reduction in subthalamic 8–35 Hz oscillatory activity correlates with clinical improvement in Parkinson’s disease. European Journal of Neuroscience. 2006;23(7):1956–1960. doi:10.1111/J.1460-9568.2006.04717.X

28. Busch JL, Kaplan J, Bahners BH, et al. Local Field Potentials Predict Motor Performance in Deep Brain Stimulation for Parkinson’s Disease. Movement Disorders. 2023;38(12):2185–2196. doi:10.1002/MDS.29626

29. Horn A, Ostwald D, Reisert M, Blankenburg F. The structural-functional connectome and the default mode network of the human brain. NeuroImage. 2014;102 Pt 1:142–151. doi:10.1016/j.neuroimage.2013.09.069

30. Fox MDM, Snyder AZA, Vincent JLJ, Corbetta MM, Van Essen DCD, Raichle MEM. The human brain is intrinsically organized into dynamic, anticorrelated functional networks. Proc Natl Acad Sci USA. 2005;102(27):9673–9678. doi:10.1073/pnas.0504136102

31. Bahners BH, Spooner RK, Hartmann CJ, Schnitzler A, Florin E. Subthalamic stimulation evoked cortical responses relate to motor performance in Parkinson’s disease. Brain Stimulation. Published online 2023. 10.1016/j.brs.2023.02.014

32. Bahners BH, Goede LL, Meyer GM, et al. Evoked response signatures explain deep brain stimulation outcomes. medRxiv. Published online October 2024:2024.10.04.24314308. doi:10.1101/2024.10.04.24314308

33. Litvak V, Eusebio A, Jha A, et al. Optimized Beamforming for Simultaneous MEG and Intracranial Local Field Potential Recordings in Deep Brain Stimulation Patients. NeuroImage. 2010;50(4):1578–1588. doi:10.1016/j.neuroimage.2009.12.115

34. Pelzer EA, Sharma A, Florin E. Data-driven MEG analysis to extract fMRI resting-state networks. Hum Brain Mapp. 2024;45(4):e26644. doi:10.1002/hbm.26644

35. Horn A, Li N, Dembek TA, et al. Lead-Dbs V2: Towards a Comprehensive Pipeline for Deep Brain Stimulation Imaging. NeuroImage. 2019;184:293–316. doi:10.1016/j.neuroimage.2018.08.068

36. Neudorfer C, Butenko K, Oxenford S, et al. Lead-DBS v3.0: Mapping deep brain stimulation effects to local anatomy and global networks. NeuroImage. 2023;268:119862. doi:10.1016/J.NEUROIMAGE.2023.119862

37. Oxenford S, Ríos AS, Hollunder B, et al. WarpDrive: Improving spatial normalization using manual refinements. Medical Image Analysis. 2024;91:103041. doi:10.1016/J.MEDIA.2023.103041

38. Husch A, V. Petersen M, Gemmar P, Goncalves J, Hertel F. PaCER - A fully automated method for electrode trajectory and contact reconstruction in deep brain stimulation. NeuroImage: Clinical. 2018;17:80–89. doi:10.1016/J.NICL.2017.10.004

39. Horn A, Kühn AA. Lead-DBS: A toolbox for deep brain stimulation electrode localizations and visualizations. NeuroImage. 2015;107:127–135. doi:10.1016/j.neuroimage.2014.12.002

40. World Medical Association Declaration of Helsinki: Ethical Principles for Medical Research Involving Human Subjects. JAMA. 2013;310(20):2191–2194. doi:10.1001/jama.2013.281053

41. Tadel F, Baillet S, Mosher JC, Pantazis D, Leahy RM. Brainstorm: A user-friendly application for MEG/EEG analysis. Computational Intelligence and Neuroscience. 2011;2011. doi:10.1155/2011/879716

42. Dahnke R, Yotter RA, Gaser C. Cortical thickness and central surface estimation. NeuroImage. 2013;65:336–348. doi:10.1016/j.neuroimage.2012.09.050

43. Veen BDV, Drongelen WV, Yuchtman M, Suzuki A. Localization of Brain Electrical Activity Via Linearly Constrained Minimum Variance Spatial Filtering. IEEE Transactions on Biomedical Engineering. 1997;44(9):867–880. doi:10.1109/10.623056

44. Brookes MJ, Hale JR, Zumer JM, et al. Measuring Functional Connectivity Using MEG: Methodology and Comparison with fcMRI. NeuroImage. 2011;56(3):1082–1104. doi:10.1016/j.neuroimage.2011.02.054

45. Brookes MJ, Woolrich M, Luckhoo H, et al. Investigating the electrophysiological basis of resting state networks using magnetoencephalography. Proceedings of the National Academy of Sciences. 2011;108(40):16783–16788. doi:10.1073/pnas.1112685108

46. Fonov V, Evans A, McKinstry R, Almli C, Collins D. Unbiased nonlinear average age-appropriate brain templates from birth to adulthood. NeuroImage. 2009;47:S102. doi:10.1016/S1053-8119(09)70884-5

47. Siddiqi SH, Schaper FLWVJ, Horn A, et al. Brain stimulation and brain lesions converge on common causal circuits in neuropsychiatric disease. Nat Hum Behav. 2021;5(12):1707–1716. doi:10.1038/s41562-021-01161-1

48. Hindriks R, Tewarie PKB. Dissociation between phase and power correlation networks in the human brain is driven by co-occurrent bursts. Commun Biol. 2023;6(1):286. doi:10.1038/s42003-023-04648-x

49. Fries P. A mechanism for cognitive dynamics: neuronal communication through neuronal coherence. Trends Cogn Sci. 2005;9(10):474–480. doi:10.1016/j.tics.2005.08.011

50. Schneider M, Broggini AC, Dann B, et al. A mechanism for inter-areal coherence through communication based on connectivity and oscillatory power. Neuron. 2021;109(24):4050–4067.e12. doi:10.1016/j.neuron.2021.09.037

51. Horn A, Neumann W, Degen K, Schneider G, Kühn AA. Toward an electrophysiological “sweet spot” for deep brain stimulation in the subthalamic nucleus. Human Brain Mapping. 2017;38(7):3377–3390. doi:10.1002/hbm.23594

52. Hollunder B, Ostrem JL, Sahin IA, et al. Mapping Dysfunctional Circuits in the Frontal Cortex Using Deep Brain Stimulation. Neurology; 2023. doi:10.1101/2023.03.07.23286766

53. Amunts K, Lepage C, Borgeat L, et al. BigBrain: An ultrahigh-resolution 3D human brain model. Science. 2013;340(6139):1472-1475. doi:10.1126/science.1235381

54. Ewert S, Plettig P, Li N, et al. Toward defining deep brain stimulation targets in MNI space: A subcortical atlas based on multimodal MRI, histology and structural connectivity. NeuroImage. 2018;170:271–282. 10.1016/j.neuroimage.2017.05.015

55. Hnazaee MF, Sure M, O’Neill GC, et al. Combining magnetoencephalography with telemetric streaming of intracranial recordings and deep brain stimulation—A feasibility study. Imaging Neuroscience. 2023;1:1–22. doi:10.1162/imag_a_00029

56. Hutchison WD, Dostrovsky JO, Walters JR, et al. Neuronal oscillations in the basal ganglia and movement disorders: evidence from whole animal and human recordings. J Neurosci. 2004;24(42):9240–9243. doi:10.1523/JNEUROSCI.3366-04.2004

57. Hirschmann J, Hartmann CJ, Butz M, et al. A direct relationship between oscillatory subthalamic nucleus–cortex coupling and rest tremor in Parkinson’s disease. Brain. 2013;136(12):3659–3670. doi:10.1093/BRAIN/AWT271

58. Hirschmann J, Steina A, Vesper J, Florin E, Schnitzler A. Neuronal oscillations predict deep brain stimulation outcome in Parkinson’s disease. Brain Stimulation. 2022;0(0). doi:10.1016/J.BRS.2022.05.008

59. Siddiqi SH, Kording KP, Parvizi J, Fox MD. Causal mapping of human brain function. Nat Rev Neurosci. 2022;23(6):361–375. doi:10.1038/s41583-022-00583-8

60. Ganos C, Al-Fatly B, Fischer JF, et al. A neural network for tics: insights from causal brain lesions and deep brain stimulation. Brain. 2022;145(12):4385–4397. doi:10.1093/brain/awac009

61. Al-Fatly B, Ewert S, Kübler D, Kroneberg D, Horn A, Kühn AA. Connectivity profile of thalamic deep brain stimulation to effectively treat essential tremor. Brain. 2019;142(10):3086–3098. doi:10.1093/brain/awz236

62. Fox MD, Buckner RL, Liu H, Chakravarty MM, Lozano AM, Pascual-Leone A. Resting-state networks link invasive and noninvasive brain stimulation across diverse psychiatric and neurological diseases. Proceedings of the National Academy of Sciences. 2014;111(41):E4367–75. doi:10.1073/pnas.1405003111

63. Fox MD, Buckner RL, White MP, Greicius MD, Pascual-Leone A. Efficacy of transcranial magnetic stimulation targets for depression is related to intrinsic functional connectivity with the subgenual cingulate. Biol Psychiatry. 2012;72(7):595–603. doi:10.1016/j.biopsych.2012.04.028

64. Weigand A, Horn A, Caballero R, et al. Prospective Validation That Subgenual Connectivity Predicts Antidepressant Efficacy of Transcranial Magnetic Stimulation Sites. Biol Psychiatry. 2018;84(1):28–37. doi:10.1016/j.biopsych.2017.10.028

65. Cole EJ, Stimpson KH, Bentzley BS, et al. Stanford Accelerated Intelligent Neuromodulation Therapy for Treatment-Resistant Depression. Am J Psychiatry. Published online April 7, 2020:appiajp201919070720. doi:10.1176/appi.ajp.2019.19070720

